# Determinants of HIV Testing Uptake Among Adolescent Girls and Young Women in Mainland Tanzania: A Stratified Analysis of the 2016/17 and 2022/2023 National Surveys

**DOI:** 10.64898/2026.02.12.26346133

**Authors:** Deogratius W. Kinoko, Anthony C. Kavindi, Paschal Yuda, Jovin R. Tibenderana, Ahmed Y. Nyaki, Sia E. Msuya, Michael J. Mahande

## Abstract

**Background:** Adolescent girls and young women (AGYW) are disproportionately vulnerable to HIV. Despite expanded HIV testing services (HTS), the majority of AGYW remain unaware of their HIV status. This study aimed to assess determinants of HIV testing uptake among AGYW in mainland Tanzania before and after stratifying by age group (15–19 and 20–24 years) using data from three national surveys conducted over time.

**Methodology:** A cross-sectional secondary data analysis was conducted using data from the Tanzania HIV Impact Surveys (2016/17 and 2022/23), obtained from the Population-based HIV Impact Assessment on 23/04/2025. Data analysis was performed using STATA version 17. Modified Poisson regression models were used to identify factors associated with HIV testing uptake before and after stratifying by age group (15–19 and 20–24 years). Results were presented using the adjusted prevalence ratio (APR) with a 95% confidence interval.

**Results:** HIV testing uptake among adolescents remained 40% in the years 2016/17 and 2022/23, while it increased from 86% to 90% among young women, respectively. Key factors consistently associated with higher prevalence of HIV testing uptake included being in a union, cohabiting, or formerly married; having secondary or higher education levels; and a history of sexually transmitted infections (STIs).

**Conclusion:** HIV testing uptake among AGYW in Tanzania has improved over time, with significant disparities between adolescents and young women. These findings highlight the need for age-specific strategies, intensifying adolescent-focused interventions while sustaining efforts among young women and reinforcing integrated reproductive health and HIV services.

## Background

Globally, HIV/AIDS persists as a major public health issue, with sub-Saharan Africa (SSA) experiencing the highest burden(1). In 2023, there were an estimated 1.9 million adolescent girls and young women aged 15–24 years living with HIV(1). Nearly 4,000 AGYW each week acquire HIV globally, and approximately 3,100 of these infections occur in SSA(1). In Tanzania, AGYW face significantly higher HIV incidence(0.33%) compared to their male peers(0.00%)(2).

Although national HIV testing services have expanded, testing uptake among AGYW remains suboptimal in Tanzania, with 40.5% of HIV-positive AGYW still unaware of their status in 2022/23, falling short of the first UNAIDS’ 95–95–95 targets(1,2). Previous studies have revealed that approximately 44-66% of new HIV infections are from HIV-infected persons who are unaware of their HIV status(3).

Increasing uptake of HIV testing and counselling and decreasing the number of undiagnosed people is identified as a priority area for HIV prevention(4). Despite ongoing initiatives like PITC, CITC, HIV self-testing, and DREAMS(5,6). Barriers such as stigma, limited youth-friendly services, and age-related legal constraints hinder testing, particularly among adolescents aged 15–19 years(4,7).

A national survey conducted in Uganda reported ever testing among adolescent girls declined from 53.8% in 2016/17 to 48.7% in 2020/21, while among young women increased from 91.9% to 93.0% respectively(8). Contrary to the national survey conducted in Tanzania that indicated ever tested for HIV among adolescent girls rose from 34.4% in 2011/12 and 38.2% in 2022/23, while among young women rose from78.4% to 89.3% respectively(2).

According to the study done in Zimbabwe, Malawi, and Tanzania, factors associated with higher HIV testing among AGYW included being aged 20-24years, living in an urban area, being married, having primary, secondary, or higher education, and having a history of pregnancy (9–12).

Due to the evidence from previous studies showing that age is a significant predictor of HIV testing(11,13), and that factors such as education, health facility visits, marital status, and ANC attendance vary between adolescents (15–19 years) and young women (20–24 years), stratified analysis by age group is justified.

Our study aimed to assess determinants of HIV testing uptake among AGYW in mainland Tanzania using data from three national surveys (THIS 2016/17and THIS 2022/23), stratified by age group, to identify disparities and inform targeted strategies.

## Conceptual framework

### Conceptual framework for analysis of factors associated with HIV testing uptake among AGYW in mainland Tanzania

We adopted Andersen’s Behavioral Model of Health Services Utilization to conceptualize the relationship between various individual and contextual factors and HIV testing uptake among AGYW(14), previously applied in studies examining HIV-related behaviors (15,16). The model comprised individual, societal, and health system levels of analysis for studying health services use and its determinants(14,16).

The framework is conceptualized based on the predisposing factors, enabling factors, and need (or perceived need) factors for health care utilization. First, the level of HIV testing uptake is independently influenced by predisposing factors, which typically include individual and contextual characteristics. Second, predisposing factors are assumed to influence HIV testing uptake indirectly through enabling factors. Third, predisposing factors may also operate through both enabling and perceived/need factors to either facilitate or hinder individuals from accessing HIV testing services (**Figure 1)**

**Figure 1:**
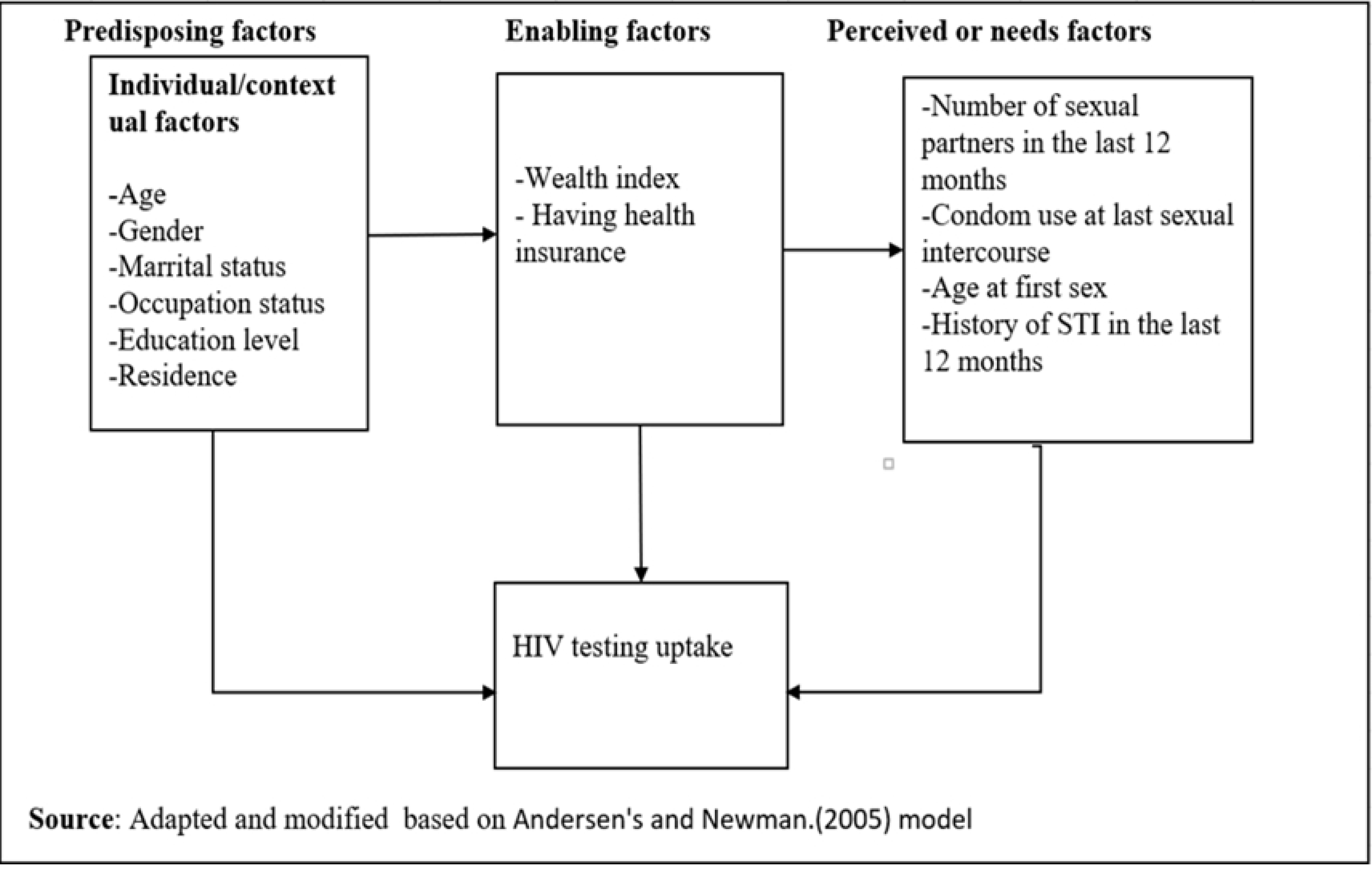
Conceptual framework for analysis of factors associated with HIV testing uptake among AGYW.

## Materials and methods

### Study design and study area

This study used a cross-sectional study design, analyzing data from the Tanzania HIV impact survey conducted between 2016/17 and 2022/23 and the dataset were accessed from the Population-based HIV Impact Assessment on 23/04/2025. Tanzania is the largest country in East Africa, spanning approximately 940,000 square kilometers, including about 60,000 square kilometers of inland water. As of 2022, the estimated AGYW population in mainland Tanzania was 5,988,919(17).

Tanzania HIV/impact surveys implemented by the National Bureau of Statistics (NBS), the Office of the Chief Government Statistician (OCGS) Zanzibar, the National AIDS, STIs and Hepatitis Control Programme (NASHCoP), and the Zanzibar Integrated HIV, Hepatitis, Tuberculosis and Leprosy Program (ZIHHTLP), with technical assistance from the US Centers for Disease Control and Prevention (CDC) and ICAP at Columbia University. The surveys are funded by the US President’s Emergency Plan for AIDS Relief (PEPFAR).

### Sampling design

The Tanzania HIV impact survey used a two-stage stratified cluster sampling design. In the first stage, enumeration areas (EAs) are selected from the national sampling frame using probability proportional to size (PPS), ensuring representativeness by region and urban-rural residence. In the second stage, a systematic sample of households is drawn from each selected EA based on an updated household listing.

### Study population and sample size

All AGYW from mainland Tanzania interviewed in the Tanzania HIV Impact Survey between 2016/17 and 2022/23 were included in the study. The number of AGYW (15–24) included in the study in the years 2016/17 and 2022/23, after applying weighting, was 6,650 and 6,064, respectively. The total weighted sample size of all AGYW included in the study was 12,714 **(Figure 2)**

**Figure 2:**
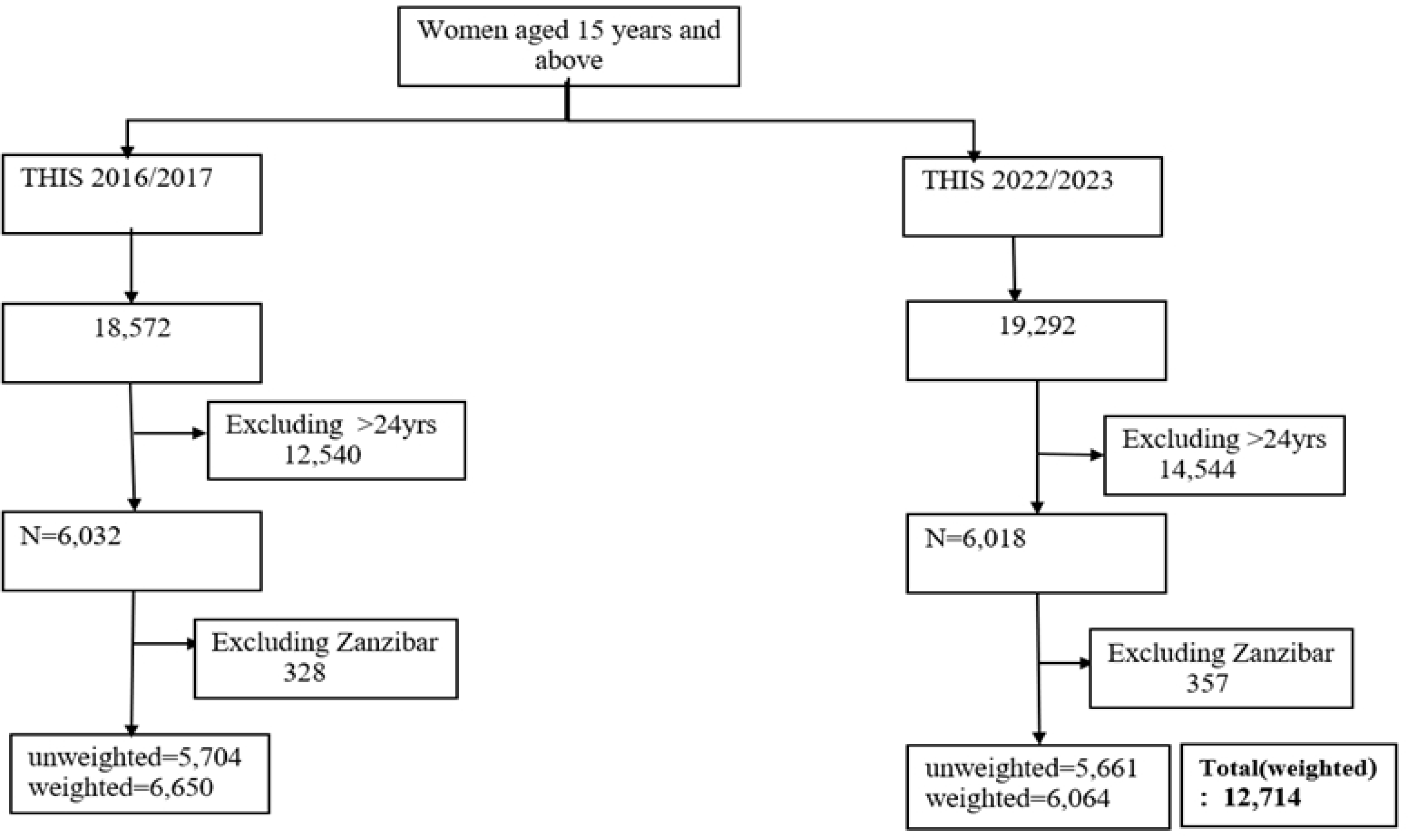
Flow chart for selection of study participants (Adolescent Girls and Young Women)

### Study variables

#### Outcome variable

The outcome of interest in this study was whether the AGYW had ever tested for HIV and received results, a binary variable if the AGYW reported “Yes” if ever been tested for HIV and received results, and “No” if not ever tested for HIV, coded as 1 “Yes” and 0 “No”.

#### Independent variables

Independent variables were classified into Predisposing factors, enabling factors, and perceived or needs factors. The predisposing factors includes: age of respondents (0=15-19 and 1=20-24years), place of residence (0=Rural, and 1=Urban), administrative zone (1=Central, 2=Lake, 3=Northen, 4= Eastern, 5=Southern West Highland, 6=Southern Highland, 7= Southern, 8=Western), marital status(0=Never in union, 1=Currently in union, 2=Cohabiting and 3=formerly in union), occupation status(0=Not employed, 1=Employed), education level(0=No education, 1=Primary education and 2=secondary education and above), exposure to TV/radio(0=No and 1=Yes). Enabling factors include: having had health insurance (0=No, and 1=Yes), Wealth index (0=poor,1=middle, and 2=Rich). Perceived or needs factor includes: age at first sex (0=<15, 1=15+), multiple sex partners in the last 12 months (0=No partner, 1=one, 2=two and above), having had STI in the last 12 months (0=No, 1=Yes), condom use in the last sex (0=No and 1=Yes). HIV results from the biomarker test (0=Negative, 1=Positive).

Zones rather than administrative regions were used in order to have consistency across the surveys. All regions belong to the same zones, and the geographical coverage of the zones has remained consistent throughout the surveys. Composition of the administrative regions in their respective zones is as follows: Eastern (Morogoro, Pwani, Dar es Salaam); Northern (Kilimanjaro, Tanga, Arusha); Lake (Mwanza, Geita, Mara, Simiyu, Shinyanga, Kagera); Central (Dodoma, Manyara, Singida); Western (Kigoma, Tabora); South West Highlands (Katavi, Rukwa, Mbeya, Songwe); Southern Highlands (Iringa, Njombe, Ruvuma); and Southern (Lindi, Mtwara).

### Data management and analysis

Data cleaning and analysis were performed using STATA version 17.0. The Variables were categorized or recategorized based on previous literature and plausibility(11,18,19). Unique ID variables merged the adult individual and biomarker datasets, and then appended across survey years to form a pooled dataset for analysis. The data analysis accounted for the complex survey design by incorporating survey weights, primary sampling units (clusters), and strata using the svyset command applied to the defined subpopulation.

Descriptive analyses are shown in frequencies and percentages for categorical variables and continuous variables using the mean with respective standard deviation to describe the AGYW characteristics. The Cochran-Armitage test for trend was used to identify significant changes in HIV testing uptake across survey years.

To determine factors associated with HIV testing uptake among AGYW, the classical logistic regression was considered. However, this approach was not used because the prevalence of HIV testing among AGYW was greater than 10% even after stratification, due to the limitation of logistic regression of overestimating odds ratios and 95%CI for outcomes with prevalence greater than 10%. Therefore, log binomial regression was then considered, but our analysis failed to converge. Finally, a modified Poisson regression model was used (20). Its functional form is in equation (i) below.

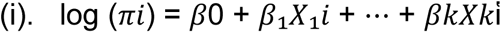

where π𝑖 is the probability of experiencing the outcome of interest for subject 𝑖

β’s is the mean of the ith subject and approximates relative ratios as exp(β).

The analysis was stratified into age groups to reflect critical differences in legal, social, developmental, and contextual factors that may influence HIV testing behaviors among adolescents and young people in mainland Tanzania. The age groups were categorized as 15–19 and 20–24 years to capture meaningful differences related to legal autonomy, maturity, and social expectations.

In Tanzania, adolescents under the age of 18 are typically required to obtain parental or guardian consent to access HIV testing services(21). This legal barrier can significantly impact access and uptake. Furthermore, developmental differences and varying levels of autonomy and risk perception between younger and older adolescents necessitate separate analysis. Combining these age groups could obscure important patterns and hinder the identification of age-specific predictors of HIV testing. Therefore, separate analyses were conducted to ensure meaningful interpretation and policy-relevant insights.

The pooled modified Poisson regression model estimated the Prevalence ratios (PR) with their corresponding 95%CI in one survey phase (2016/17-2022/23). Any variable with a P-value <0.2 in univariate analysis and a variable that is important in explaining the outcome based on the conceptual framework and previous study were included in the multivariable model. Forward stepwise regression was performed. The model had the lowest AIC, suggesting it was the most parsimonious model for my study. Statistical significance was determined at a p-value of less than 0.05. The variable was regarded as a confounder if the changes in the estimate from crude to adjusted analysis were≥10%. Also, multicollinearity between exposures was checked. The variance inflation factor (VIF<5) was considered as low collinearity.

## Results

### Background characteristics of the study participants

A total of 12,714 participants were analyzed across the two surveys, with a mean (SD) age of 19.4(±2.8). Across the two surveys (2016/17 and 2022/23), the majority of participants were aged 15–19 years, although their proportion decreased over time from 54.2% in 2016/17 to 51.4% in 2022/23. Additionally, the majority resided in rural areas, and their proportion remained constant 59.3% in the years 2016/17 and 2022/23. The majority of AGYW were never married, with the proportion remaining relatively consistent at approximately 56% throughout all survey years. Regarding education, most AGYW had attained primary education, although this proportion declined over time from 55% in 2016/17 to 48% in 2022/23. The majority of AGYW reported having one sexual partner, with proportions increasing from 51.7% in 2016/17 to 63.9% in 2022/23. Additionally, the majority of AGYW reported having had no sexually transmitted infection (STI) in the 12 months before the survey, with the proportions dropping from 87.5% in 2016/17 to 81.3% in 2022/23**(Table 1**).

**Table 1:** Background characteristics of the study participants (weighted) in mainland Tanzania (N=12,714)

| Characteristics | Survey year |  |
| --- | --- | --- |
|  | 2016/2017 (n=6,650)<br>n(%) | 2022/2023 (n=6,064)<br>n(%) |
| **Age(years)** |  |  |
| 15-19 | 3,603(54.2) | 3,115(51.4) |
| 20-24 | 3,047(45.8) | 2,949(48.6) |
| **Mean (SD)** | 19.4(2.8) | 19.6(2.8) |
| **Residence** |  |  |
| Rural | 3,943(59.3) | 3,600(59.4) |
| Urban | 2,707(40.7) | 2,464(40.6) |
| **Zone** |  |  |
| Central | 698(10.5) | 542(8.9) |
| Lake | 1,129(27.5) | 1,928(31.8) |
| Northern | 750(11.3) | 640(10.6) |
| Eastern | 1,309(19.7) | 1,234(20.4) |
| South West Highland | 813(12.2) | 594(9.8) |
| Southern Highland | 489(7.4) | 322(5.3) |
| Southern | 190(2.9) | 226(3.7) |
| Western | 571(8.6) | 577(9.5) |
| **Marital status** |  |  |
| Never in union | 3,705(55.7) | 3,383(55.8) |
| Currently in a union | 1,875(28.2) | 1,839(30.3) |
| Cohabiting | 706(10.6) | 531(8.8) |
| Formerly in union | 364(5.5) | 312(5.1) |
| **Occupation status** |  |  |
| Not employed | 4,952(74.5) | 4,323(71.3) |
| Employed | 1,699(25.5) | 1,741(28.7) |
| **Education Level** |  |  |
| No education | 551(8.3) | 466(7.7) |
| Primary education | 3,655(55) | 2,913(48) |
| Secondary/Higher | 2,445(36.7) | 2,685(44.3) |

<sup>a</sup>-Sex debut 2016/17(n=4,677); <sup>b</sup>-Multiple sexual partners 2016/17(n=6,612);
<sup>c</sup>-Condom use 2011/12(n=2,516),2016/17(n=6,089),2022/23(n=5,804); <sup>e</sup>-Had STI in last 12months 2016/17(n=4,677), 2022/23(n=5,469)

**Table 1 Continued: Background characteristics of the study participants(weighted) in mainland Tanzania (N=12,714)**
| Characteristics | Survey year |  |
| --- | --- | --- |
|  | 2016/17(n=6650)<br>n(%) | 2022/23(n=6064)<br>n(%) |
| **Wealth index** |  |  |
| Poor | 2,417(36.3) | 2,472(40.8) |
| Middle | 1,389(20.9) | 1,229(20.3) |
| Rich | 2,845(42.8) | 2,363(38.9) |
| **Exposure to Radio/TV** |  |  |
| No | 2,417(36.3) | 1,842(30.4) |
| Yes | 4,234(63.7) | 4,222(69.6) |
| **Had health insurance** |  |  |
| No | 5,971(89.8) | 4,817(79.4) |
| Yes | 680(10.2) | 1,247(20.6) |
| **Age at first sex <sup>a</sup>** |  |  |
| <15 | 615(13.2) | 351(8.7) |
| 15+ | 4,062(86.8) | 3,665(91.3) |
| **Multiple sexual partners <sup>b</sup>** |  |  |
| No partner | 583(8.8) | 298(5.0) |
| One | 3,421(51.7) | 3,642(60.7) |
| Two and above | 2,608(39.5) | 2,058(34.3) |
| **Condom use in last sex <sup>c</sup>** |  |  |
| No | 3,197(52.5) | 3,327(57.3) |
| Yes | 2892(47.5) | 2,477(42.7) |
| **Had STI in the last 12months<sup>e</sup>** |  |  |
| No | 4,091(87.5) | 4,447(81.3) |
| Yes | 586(12.5) | 1,022(18.7) |
| **HIV results from the biomarker test.** |  |  |
| Positive | 141(2.1) | 78(1.3) |
| Negative | 6,509(97.9) | 5,986(98.7) |

<sup>a</sup>-Sex debut 2016/17(n=4,677); <sup>b</sup>-Multiple sexual partners 2016/17(n=6,612);
<sup>c</sup>-Condom use 2011/12(n=2,516), 2016/17(n=6,089), 2022/23(n=5,804); <sup>e</sup>-Had STI in last 12months 2016/17(n=4,677), 2022/23(n=5,469)

### Trends of HIV testing uptake among adolescent girls and young women

Trends in HIV testing among AGYW (15-24years) who had ever been tested and received results, the proportion increased from 61% in 2016/17 and 64% in 2022/23. When stratified by age, HIV testing uptake among adolescent girls aged 15-19 years remained 40% in both 2016/17 and 2022/23 surveys. In contrast, among young women aged 20-24 years, HIV testing increased from 86% to 90%, respectively (**Figure 3**).

**Figure 3:**
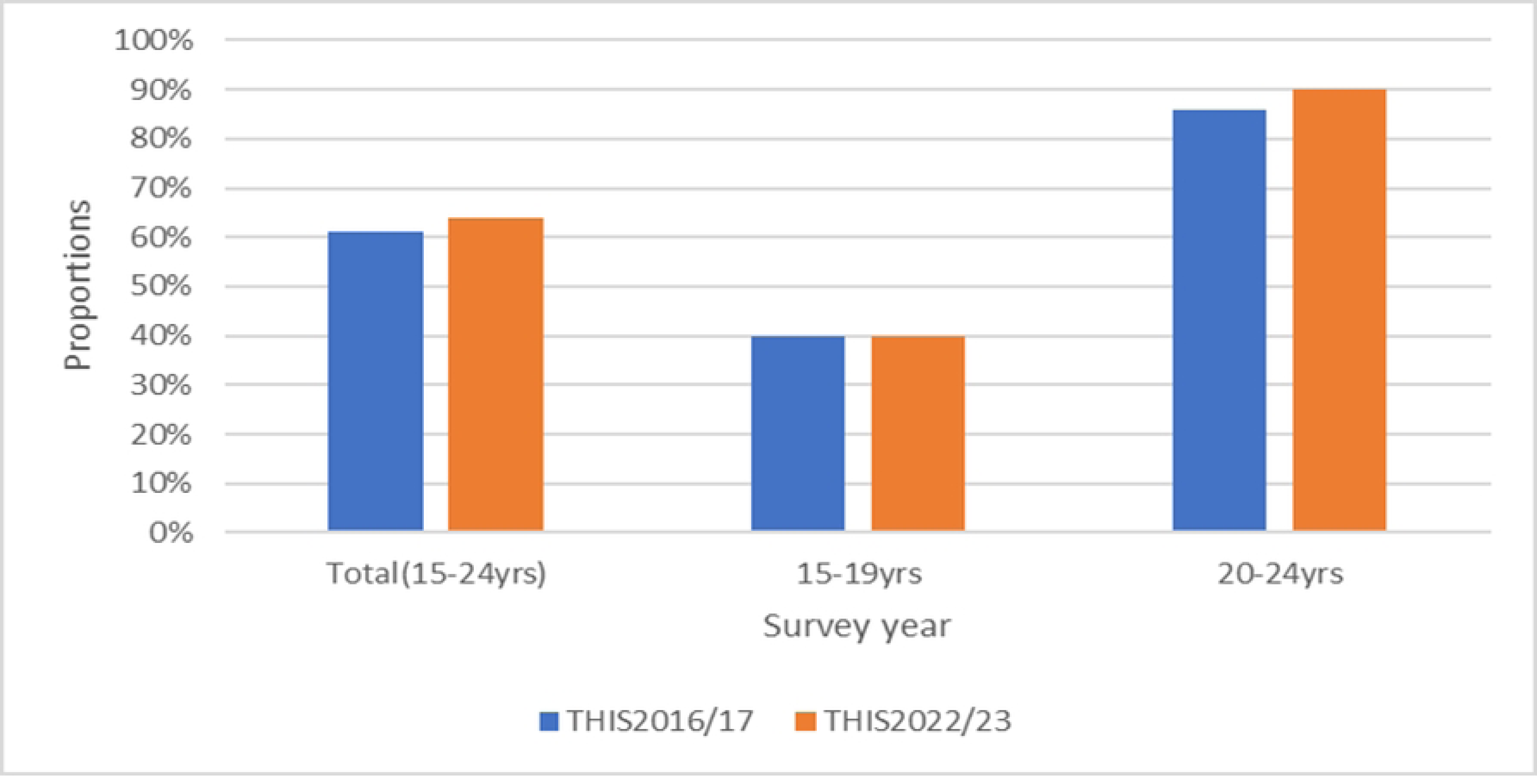
Trends in ever tested for HIV and received results among AGYW in mainland Tanzania between THIS2016/17, and 2022/23.

### Distribution of HIV testing uptake according to selected characteristics

Our results from two surveys (2016/17 to 2022/23) found a significant increase in percentages of HIV testing across age groups, marital status, employment status, and condom use in the last sex (P <0.001). Among AGYW aged 20-24 years increased from 86.2% to 89.8% (a 3.6%-point increase). Among AGYW who were currently in union increased from 86.2% to 93.1% (a 6.9%-point increase). Among employed rose from 70.4% to 74.7% (a 4.3%-point increase), and among AGYW who did not use a condom in the last sex increased from 80.1% to 87.6% (a 7.5%-point increase) (**Table 2**).

**Table 2:** Trends of ever tested for HIV and received results by selected characteristics of study participants (Chi-square test) in THIS2016/17, and THIS2022/23 (N=12,714)

| variables | Ever tested for HIV |  | % Differences |
| --- | --- | --- | --- |
|  | 2016/17(%) | 2022/23(%) | 2016/17-2022/23 |
| **Age (years)** |  |  |  |
| 15-19 | 39.9*** | 40.0*** | 0.1 |
| 20-24 | 86.2 | 89.8 | 3.6 |
| **Residence** |  |  |  |
| Rural | 59.6 | 63.6 | 4.0 |
| Urban | 63.3 | 65.0 | 1.7 |
| **Marital status** |  |  |  |
| Never in union | 40.7*** | 41.3*** | 0.6 |
| Currently in a union | 86.2 | 93.1 | 6.9 |
| Cohabiting | 87.4 | 92.6 | 5.2 |
| Formerly in union | 87.8 | 94.1 | 6.3 |
| **Occupation status** |  |  |  |
| Not employed | 57.9*** | 60.0*** | 2.1 |
| Employed | 70.4 | 74.7 | 4.3 |
| **Education level** |  |  |  |
| No education | 64.7** | 76.2*** | 11.5 |
| Primary education | 63.1 | 67.8 | 4.7 |
| Secondary/higher | 57.3 | 58.2 | 2.0 |
| **Zone** |  |  |  |
| Central | 61.3** | 59.1** | 8.2 |
| Lake | 58.9 | 62.2 | 4.3 |
| Northern | 53.3 | 60.9 | 15.3 |
| Eastern | 62.1 | 66.2 | 2.5 |
| South West Highland | 63.5 | 69.7 | 2.9 |
| Southern Highland | 71.0 | 73.9 | 8.6 |
| Southern | 68.2 | 60.2 | 1.9 |
| WeIndex | 61.8 | 65.7 | 4.2 |
| **Household Wealth Index** |  |  |  |
| Poor | 59.6 | 64.4 | 7.5 |
| Middle | 62.0 | 61.6 | 4.5 |
| Rich | 61.9 | 65.4 | 4.3 |

**Table 2 continued: Trends of ever tested for HIV and received results by selected characteristics of study participants (Chi-square test) in THIS2016/17, and THIS2022/23 (N=12,714)**
| Variables | Ever tested for HIV |  | % Differences |
| --- | --- | --- | --- |
|  | 2016/17 (%) | 2022/23 (%) | 2016/17-2022/23 |
| **Exposure to Radio/TV** |  |  |  |
| No | 59.6 | 68.6** | 9.0 |
| Yes | 61.9 | 62.3 | 0.4 |
| **Health insurance** |  |  |  |
| No | 60.6 | 64.1 | 3.5 |
| Yes | 65.2 | 64.8 | -0.4 |
| **Sexual debut** |  |  |  |
| <15 | 67.5*** | 79.2** | 11.7 |
| 15+ | 80.2 | 85.9 | 5.7 |
| **Multiple sexual partners** |  |  |  |
| No partner | 75.6*** | 61.3 | -14.3 |
| One | 80.0 | 84.2 | 4.2 |
| Two and above | 18.2 | 28.9 | 10.7 |
| **Condom use in the last sex** |  |  |  |
| No | 80.1*** | 87.6*** | 7.5 |
| Yes | 37.0 | 32.7 | -4.3 |
| **Had STI in the past 12months** |  |  |  |
| No | 78.8 | 53.4*** | -33 |
| Yes | 76.8 | 94.7 | 17.9 |
| **HIV results from a biomarker test** |  |  |  |
| Negative | 60.6*** | 63.9*** | 3.3 |
| Positive | 84.6 | 91.1 | 6.5 |

Decline in proportion of HIV testing; \*Significant at P<0.05; \*\*Significant at P<0.01; \*\*\*Significant at P<0.001

### Proportion of HIV testing uptake among AGYW by Regions

Between 2016/17 and 2022/23, half of the regions in mainland Tanzania recorded a significant increase in HIV testing among AGYW beyond the national average (63%). These regions are Tanga, Katavi, Mara, Dar es-Salaam, Njombe, Rukwa, Shinyanga, Mbeya, Tabora, Morogoro, Ruvuma, Songwe, and Iringa. These regional variations highlight both progress and gaps in HIV testing efforts, underscoring the need for targeted interventions in areas where proportions have declined **(Figure 4).**

**Figure 4:**
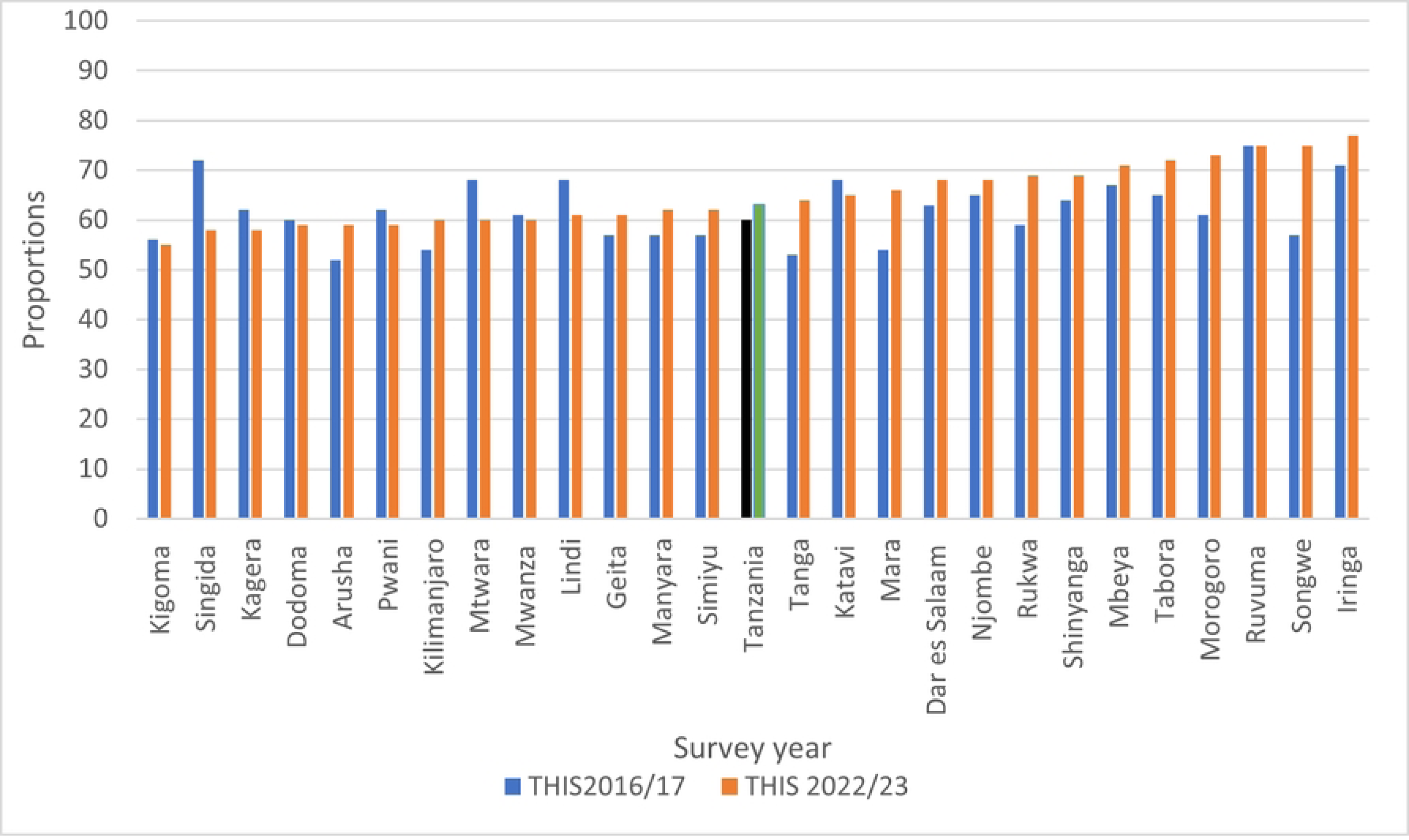
Trends in ever tested for HIV and received results among AGYW in mainland Tanzania by regions 2016/17, and 2022/23.

### Factors Associated with HIV Testing Uptake among AGYW in Mainland Tanzania (2016/17–2022/23)

**Table 3** presents the results from pooled multivariable modified Poisson regression models assessing factors associated with HIV testing uptake among AGYW in mainland Tanzania. HIV testing prevalence was 31% higher among AGYW aged 20–24 years compared to those aged 15–19 (APR: 1.31, 95% CI: 1.26–1.37, p < 0.001), indicating age as a strong predictor of testing uptake. However, a 42% reduction in the adjusted prevalence ratio suggests that the effect of age is partially confounded by other variables such as marital status and sexual activity.

**Table 3:** Pooled multivariable Poisson regression on factors associated with HIV testing among AGYW in mainland Tanzania using data from THIS 2016/17 and 2022/23 (N=12,714)

| Variables | CPR (95%CI) | P-value | APR (95%CI) | p-value |
| --- | --- | --- | --- | --- |
| **Age(years)** |  |  |  |  |
| 15-19 | 1 |  | 1 |  |
| 20-24 | 2.26(2.16-2.37) | <0.001 | 1.31(1.26-1.37) | <0.001 |
| **Residence** |  |  |  |  |
| Rural | 1 |  | 1 |  |
| Urban | 1.07(1.02-1.12) | 0.003 | 1.04(0.99-1.07) | 0.052 |
| **Zone** |  |  |  |  |
| Central | 1 |  | 1 |  |
| Lake | 0.99(0.93-1.05) | 0.996 | 0.96(0.88-1.06) | 0.872 |
| Northern | 0.99(0.85-1.15) | 0.817 | 1.00(0.87-1.17) | 0.660 |
| Eastern | 1.08(0.91-1.13) | 0.054 | 0.94(0.84-1.04) | 0.584 |
| SouthW highland | 0.99(0.88-1.17) | 0.952 | 0.99(0.88-1.11) | 0.992 |
| Southern highland | 1.01(0.88-1.17) | 0.876 | 1.01(0.89-1.14) | 0.763 |
| Southern | 1.13(0.99-1.29) | 0.222 | 1.09(0.97-1.23) | 0.717 |
| Western | 0.99(0.88-1.13) | 0.180 | 1.00(0.89-1.11) | 0.200 |
| **Marital status** |  |  |  |  |
| Never in union | 1 |  | 1 |  |
| Currently union | 2.1(2.00-2.27) | <0.001 | 1.29(1.20-1.28) | <0.001 |
| Cohabiting | 2.21(2.08-2.42) | <0.001 | 1.25(1.19-1.31) | <0.001 |
| Formerly in union | 2.35(2.14-2.53) | <0.001 | 1.20(1.20-1.34) | <0.001 |
| **Occupation status** |  |  |  |  |
| Not employed | 1 |  | 1 |  |
| Employed | 1.18(1.12-1.23) | <0.001 | 0.97(0.95-1.01) | 0.192 |
| **Education Level** |  |  |  |  |
| No education | 1 |  | 1 |  |
| Primary | 0.97(0.89-1.05) | 0.456 | 1.11(1.03-1.19) | 0.003 |
| Secondary/higher | 0.89(0.82-0.98) | 0.021 | 1.16(1.07-1.26) | <0.001 |
| **Wealth index** |  |  |  |  |
| Poor | 1 |  | 1 |  |
| Middle | 0.96(0.90-1.02) | 0.183 | 0.99(0.98-1.11) | 0.982 |
| Rich | 1.02(0.97-1.07) | 0.444 | 1.01(0.97-1.06) | 0.564 |

**Table 3 continued: pooled multivariable Poisson regression on factors associated with HIV testing among AGYW in mainland Tanzania using data from THIS 2016/17 and 2022/23 (N=12,714)**
| Variables | CPR (95%CI) | p-value | APR (95%CI) | p-value |
| --- | --- | --- | --- | --- |
| **Exposure to radio/TV** |  |  |  |  |
| No | 1 |  | 1 |  |
| Yes | 0.95(0.91-0.99) | 0.023 | 1.00(0.97-1.03) | 0.906 |
| **Had health insurance** |  |  |  |  |
| No | 1 |  | 1 |  |
| Yes | 1.07(1.01-1.14) | 0.029 | 0.95(0.88-1.020) | 0.867 |
| **Sexual debut** |  |  |  |  |
| <15 | 1 |  | 1 |  |
| 15+ | 2.05(1.86-2.26) | <0.001 | 1.03(0.94-1.14) | 0.813 |
| **Multiple sex partners** |  |  |  |  |
| No partner | 1 |  | 1 |  |
| One | 2.30(2.11-2.50) | <0.001 | 1.30(1.00-1.9) | 0.030 |
| Two or more | 0.91(0.81-1.01) |  | 0.85(0.77-1.05) | 0.107 |
| **Condom use** |  |  |  |  |
| No | 1 |  | 1 |  |
| Yes | 0.49(0.47-0.51) | <0.001 | 1.04(1.00-1.07) | 0.046 |
| **Had an STI in the last 12 months** |  |  |  |  |
| No | 1 |  | 1 |  |
| Yes | 1.69(1.64-1.76) | <0.001 | 1.22(1.18-1.26) | <0.001 |
| **HIV results from the biomarker test** |  |  |  |  |
| Negative | 1 |  | 1 |  |
| Positive | 1.37(1.27-1.49) | <0.001 | 1.06(0.91-1.22) | 0.238 |

1-reference group: CPR-Crude Prevalence Ratio; APR-Adjusted Prevalence Ratio

Being in a union was consistently associated with higher HIV testing uptake. AGYW who were currently married, cohabiting, or formerly in union had 29%, 25%, and 20% higher prevalence of testing, respectively, compared to those never in union. Yet, after adjusting for confounders, the strength of these associations decreased by 39–49%, indicating that marital status likely acts as a proxy for other underlying factors, particularly age and sexual exposure. Education also played a significant role. AGYW with at least secondary education had a 16% higher prevalence of HIV testing compared to those with no formal education (APR: 1.16, 95% CI: 1.07–1.26, p < 0.001), reflecting the positive impact of educational attainment on health-seeking behavior. Recent STI history emerged as a strong independent predictor. AGYW who reported having had an STI in the past 12 months had a 22% higher likelihood of undergoing HIV testing (APR: 1.22, 95% CI: 1.18–1.26, p < 0.001), underscoring how perceived risk increases demand for testing.

Sexual behavior also influenced testing. AGYW with one sexual partner had 30% higher HIV testing prevalence than those who had never had a partner (APR: 1.30, 95% CI: 1.00–1.90, p = 0.030). However, this association weakened by 44% after adjustment, suggesting it was likely confounded by other risk indicators such as age, marital status, and STI experience.

Structural and contextual variables, including geographic zone, urban/rural residence, employment status, household wealth index, exposure to TV or radio, and HIV biomarker status, were not significantly associated with HIV testing in the adjusted models. This suggests that individual-level characteristics (e.g., age, education, sexual activity) may exert a greater influence on testing behaviors than broader environmental or structural determinants. It is also possible that the effects of structural factors are mediated through individual characteristics, or that limited variability and measurement constraints reduced the power to detect significant associations.

### Factors associated with HIV testing among AGYW in mainland Tanzania stratified by age using data from THIS 2016/17 and 2022/23

When examining factors associated with HIV testing uptake among AGYW in mainland Tanzania stratified by age groups (adolescents aged 15–19 and young women aged 20–24) using data from the 2016/17 and 2022/23 THIS surveys, similar predictors were identified across both groups. These included being in a union (currently married, cohabiting, or formerly in a union), attaining secondary education or higher, and having had a sexually transmitted infection (STI) within the past year (**Table 4)**. The consistency of these associations suggests common individual-level drivers of HIV testing across the adolescent and young adult age spectrum.

**Table 4:**
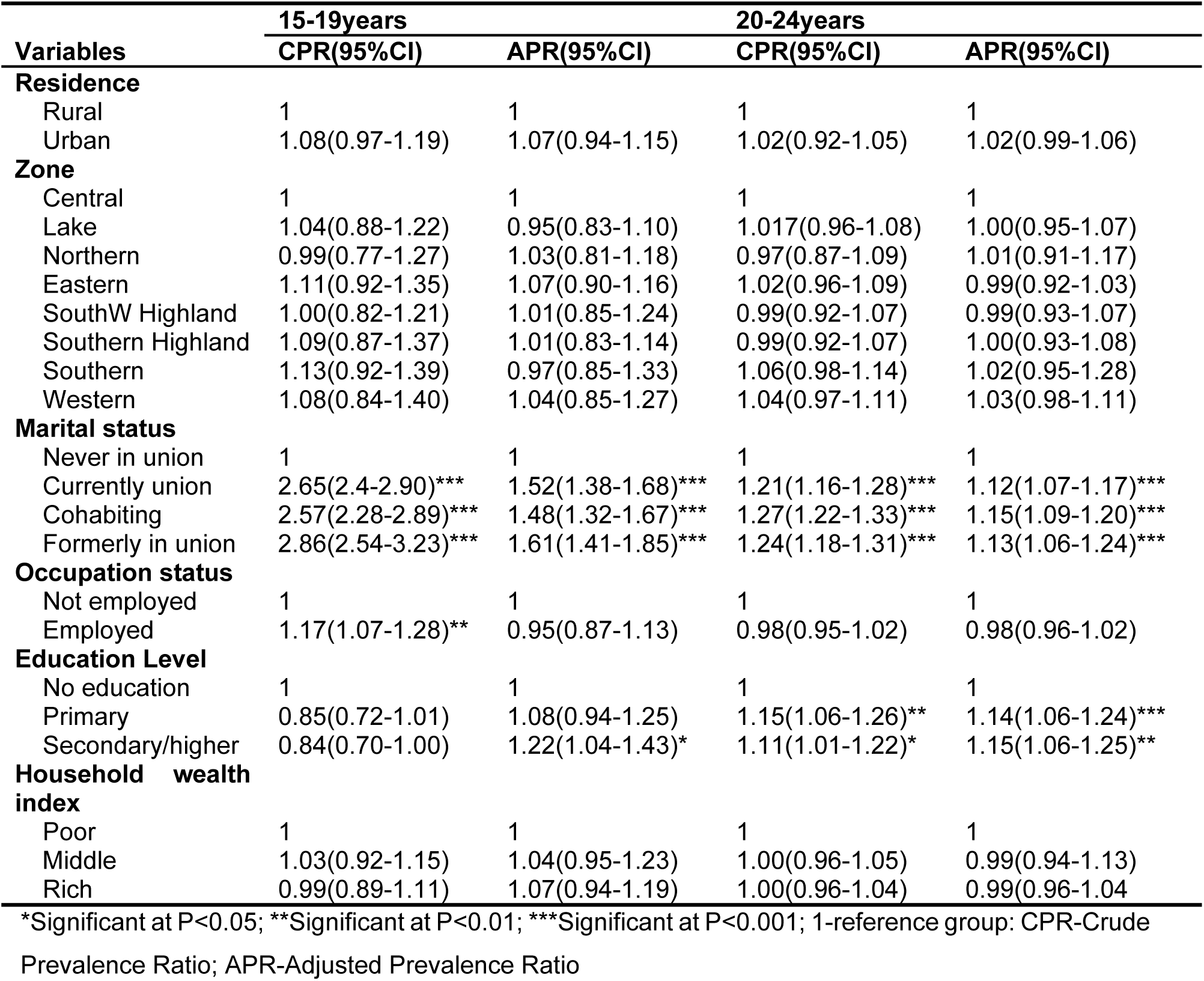
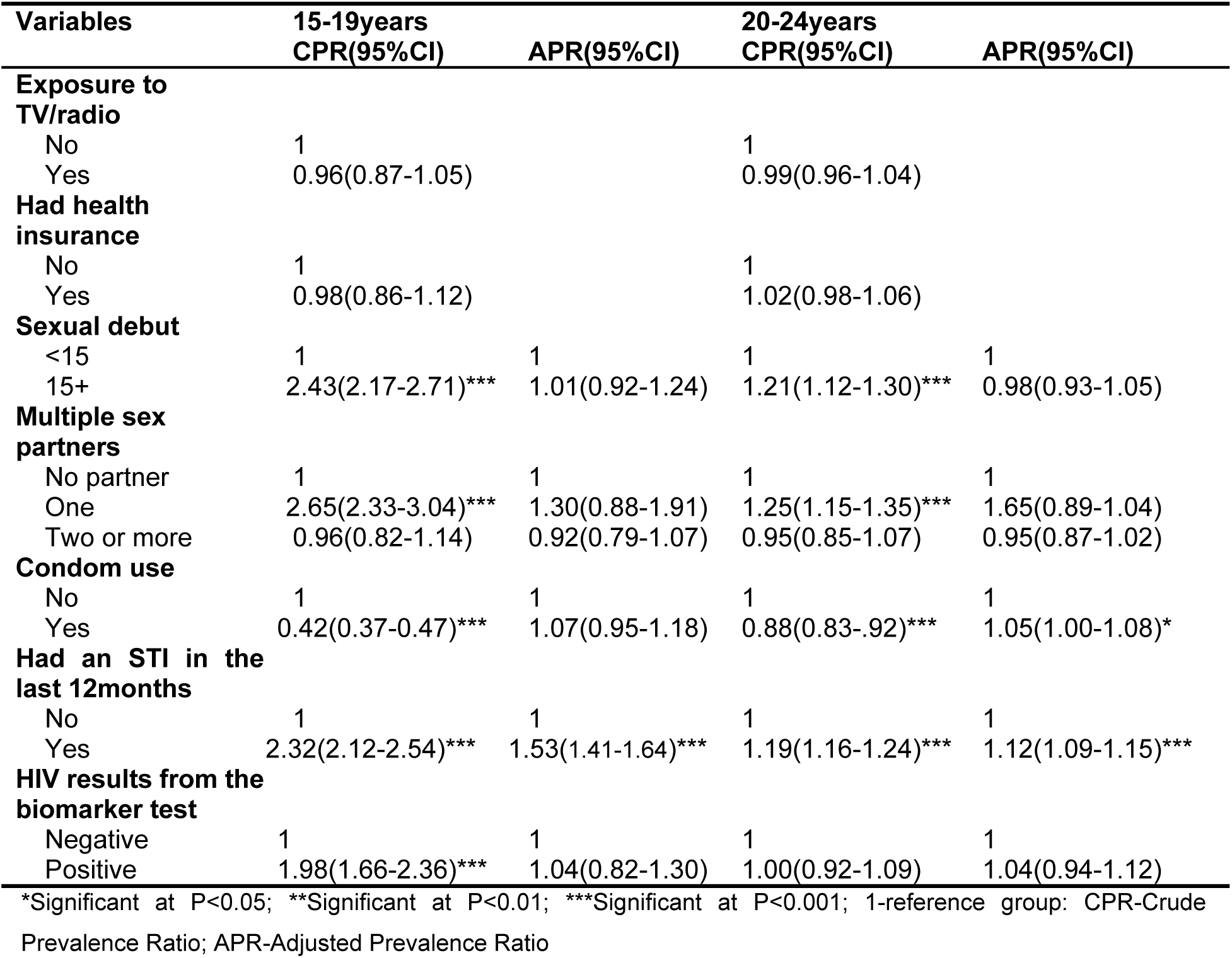
Pooled multivariable modified Poisson regression on factors associated with HIV testing among AGYW in mainland Tanzania stratified by age using data from THIS 2016/17 and 2022/23 (N=12,714)

However, after adjusting for potential confounders, the association between marital status and HIV testing uptake among adolescents aged 15–19 was notably attenuated. Prevalence ratios dropped by 43% for those currently in union, 42% for those cohabiting, and 44% for those formerly in union. This suggests that marital status in this age group may not independently influence HIV testing, but rather acts as a proxy for other factors such as age, sexual debut, or STI risk, each of which more directly motivates testing.

In contrast, the corresponding reductions among young women aged 20–24 were modest (7% to 9%), indicating that marital status remains a more stable, independent predictor of HIV testing in this older cohort. This could reflect greater autonomy, increased exposure to health services, or higher perceived HIV risk post-marriage.

Our findings underscore the importance of considering age-specific dynamics when designing HIV testing interventions. For adolescents, strategies may need to go beyond marital status and directly address sexual health education, STI prevention, and accessibility of youth-friendly HIV testing services.

## Discussion

The findings from this study indicate that the overall proportion of HIV testing uptake among AGYW increased from 61% in 2016/17 to 64% in 2022/23. HIV testing uptake was positively associated with being aged 20–24 years, currently or formerly in union, cohabiting, having had a sexually transmitted infection (STI), and having attained secondary education or higher.

Among young women aged 20–24 years, HIV testing coverage improved significantly from 86% in 2016/17 to 90% in 2022/23. In contrast, the uptake among adolescent girls aged 15–19 years remained 40% in both survey years. These results are consistent with trends reported in other sub-Saharan African countries. For example, in Lesotho, HIV testing among adolescents increased from 26.5% in 2010/11 to 47.9% in 2015(10), a greater relative improvement than observed in Tanzania. The persistently low uptake among adolescents in Tanzania may reflect structural and policy barriers, such as the requirement for parental consent for individuals under 18 years to access HIV testing services(21). In contrast, countries like Lesotho permit self-consent from age 12, potentially facilitating earlier engagement with HIV services(22). Higher testing rates among young women compared to adolescents may be partially explained by their greater engagement with healthcare services, particularly through antenatal care (ANC) and maternal health programs. Tanzanian national guidelines recommend provider-initiated HIV testing during ANC visits, which may increase testing opportunities for women in this age group.

Our study examined the factors associated with HIV testing uptake among adolescent girls and young women (AGYW) in mainland Tanzania based on THIS (2016/17 and 2022/23). Our study revealed that being AGYW aged 20-24 years, currently in union, cohabiting, and formerly in union, having attained secondary education or higher, and having had an STI were the factors consistently and positively independently associated with HIV testing among AGYW. After stratifying by age group, both adolescents(15-19years) and young women (20-24 years) exhibited similar factors associated with HIV testing as observed in the overall analysis.

This study revealed that HIV testing uptake changes with, women aged 20–24 years had a higher prevalence of getting tested for HIV as to with women aged 15–19 years. This finding is consistent with previous studies(9–12). This could be explained by the fact that women aged 20-24 years of age are more likely to be sexually active, married, cohabiting or pregnant which increases their exposure to routine provider-initiated HIV testing through antenatal and reproductive health services. They tend to have greater decision-making power better knowledge on HIV and lower perceived stigma than adolescents aged 15-19 years of age who usually face social and parental barriers to accessing testing, fear of judgement and underestimate their risk. Eventually, these structural, behavioral and service-related factors collectively drive higher HIV testing uptake among these women

Marital status was positively associated with HIV testing, with higher uptake observed among women who were currently in union, cohabiting, or formerly in union. These findings are aligned with previous studies in Tanzania and elsewhere (18,23). The following could be reasons for these findings, first partnered women have increased contact with health services through antenatal care, family planning and couple-based interventions where testing is routinely recommended. Being in or having been in partnership heightens perceived HIV risk related to partner fidelity and future child bearing, motivating acceptance of testing. Furthermore, healthcare providers may prioritize testing among partnered women to support prevention of mother to child transmission and partner notification while social support from partner can reduce fear, stigma and anxiety

The history of sexually transmitted infections (STIs) was also strongly associated with HIV testing, likely reflecting the integration of HIV testing into STI diagnosis and treatment services. Similar associations have been reported in Senegal and Haiti, where individuals with STI symptoms were more likely to seek or be referred for HIV testing(24). However, earlier studies in Tanzania and Zimbabwe reported no such association(9,11). This discrepancy may be due to the limited implementation or reach of integrated HIV and STI services at that time, which may have hindered routine referrals of AGYW for HIV testing.

Education level emerged as another important determinant, with AGYW who had completed secondary education or higher being significantly more likely to have ever tested for HIV. This association is consistent with findings from multiple countries, including Zambia, Burundi, and South Africa(16,25). Increased educational attainment may enhance awareness of HIV transmission, reduce stigma, and empower young women to access testing services.

Contrary to previous studies(11,15), the place of residence (rural vs. urban) was not significantly associated with HIV testing uptake in this study, even after adjusting for confounders. This may suggest that HIV testing services have become more equitably distributed across geographic areas in Tanzania, likely due to the nationwide scale-up of community-based and mobile testing programs. Additionally, place of residence may correlate with wealth, education, and access to media; adjusting for these reduces its independent association.

Our study revealed that the region(zone) was not significantly associated with HIV testing uptake. In contrast, a study conducted in Tanzania by Mahande et al. (2016). This may partly be due to small sample sizes in some regions, which limit the statistical power to detect differences. Furthermore, the uniform availability of HIV testing services across different regions may have reduced regional disparities in HIV testing.

## Strengths and Limitations

This study is the first in Tanzania to comprehensively examine determinants of HIV testing among AGYW by age strata, using robust nationally representative survey data spanning over a decade. The large sample sizes provided sufficient power for subgroup analysis.

However, the study is not without limitations. The use of secondary data introduces potential issues related to missing or unmeasured variables. Self-reported HIV testing history may be subject to recall or social desirability bias, particularly among adolescents. Additionally, certain important contextual and behavioral variables (e.g., stigma, peer influence, knowing a place for HIV testing, alcohol use, and sexual violence) were not captured in the datasets consistently, limiting the interpretation of some observed associations.

## Conclusion and Recommendations

HIV testing uptake among AGYW in mainland Tanzania has improved between 2016/72 and 2022/2023, with notable gains among young women aged 20–24 years. However, progress among adolescents remains limited. Key factors consistently associated with a higher prevalence of HIV testing include being in a union, a history of STIs, and higher educational attainment. Targeted efforts are needed to address persistent barriers and expand access to adolescent-friendly HIV testing services.

Strengthen and promote couple HIV Testing and Counselling (CHTC), particularly for AGYW currently in union or cohabiting relationships. This approach enhances mutual disclosure, reduces stigma, and promotes shared responsibility in HIV prevention and care.

Integrate HIV testing with reproductive and STI services: Programs should continue strengthening HIV testing during STI care and expand testing access through youth-friendly health services.

Strengthen school-based sexual and reproductive health (SRH) education: efforts should focus on enhancing their quality, coverage, and effectiveness. The Tanzania Education and Training Policy (ETP) 2014 Edition of 2023 outlines several key areas, including the integration of health education in primary, secondary, and higher levels, to overcome health challenges(26). Further qualitative studies are needed to explore social, cultural, and psychological barriers to HIV testing among adolescents, particularly those aged 15–19 years.

## Ethical clearance

Each survey received ethical clearance from the National Institute for Medical Research (NIMR) in Tanzania and the Zanzibar Medical Research and Ethics Committee (ZMREC), as well as approval from relevant international institutional review boards. Informed consent was obtained from all participants before data collection, with assent and parental permission required for minors. For this secondary analysis, ethical approval to conduct the study was sought from the KCMC University Research and Ethics Review Committee (KURERC) with clearance number PG 180/2024. Permission to access the datasets was granted by the Tanzania National Data Archive (TNADA) and the PHIA project website following submission of a project abstract.

## Data Availability

The data and materials used in this study are available for free and on request on the PHIA project website at https://phia.icap.columbia.edu and on the National Bureau of Statistics website at www.nbs.go.tz.

https://www.nbs.go.tz/

## Acknowledgments

We acknowledge and appreciate the academic staff of the Institute of Public Health– KCMC University for their support. The National Bureau of Statistics is conducting the surveys and providing the dataset.

## Data Availability

The data and materials used in this study are available for free and on request on the PHIA project website at http://phia.icap.columbia.edu and on the National Bureau of Statistics website at www.nbs.go.tz.

## List of abbreviations

AGYW: Adolescent Girl and Young Women
CHTC: Couple HIV Testing and Counselling
CITC: Client-Initiated HIV Testing and Counselling
DHS: Demographic Health Survey
HIV: Human Immune Deficiency Virus
HIVST: HIV Self Testing
HTS: HIV Testing Services
MoH: Ministry of Health
NASHCoP: National AIDS, STIs and Hepatitis Control Programme
PITC: Provider-Initiated HIV Testing and Counselling
STIs: Sexually Transmitted Infections
THIS: Tanzania HIV Impact Survey
UNAIDS: United Nations Programme on HIV/AIDS
VCTC: Voluntary Testing and Counselling Center
WHO: World Health Organization

